# Return to Work with Long COVID: A Rapid Review of Support and Challenges

**DOI:** 10.1101/2025.04.29.25326647

**Authors:** Sarah Daniels, Hua Wei, Damien M McElvenny, Martie van Tongeren, Donna Bramwell, Anna Coleman, Davine Forde, Ruth Wiggans

## Abstract

**Objective:** To explore existing evidence for the provision of support for return to work (RTW) in long COVID (LC) patients, and the barriers and facilitators to taking up this support.

**Methods:** A rapid review was reported according to the Preferred Reporting Items for Systematic Reviews and Meta-Analyses (PRISMA) approach. Searches were completed in June 2024 and included MEDLINE, Embase, American Psychological Association (APA) PsycINFO, evidence based medicine (EBM) Reviews (including the Cochrane Central Register of Controlled Trials and Cochrane Database of Systematic Reviews), Health Management Information Consortium, Web of Science and Google Scholar. This review included studies on LC symptoms lasting over 12 weeks, focusing on 1) non-workplace- and workplace-based support for RTW in LC patients, and/or 2) barriers and facilitators to RTW in LC patients. A quality assessment was conducted using the JBI Systematic Reviews critical appraisal tool. The data were summarised in tabular format and a narrative summary. This study was pre-registered (PROSPERO-ID: CRD42023478126).

**Results:** Twenty-five studies were included. While many studies demonstrated rigorous methodologies and low risk of bias levels, some had high and medium risk levels. Non-workplace-based support was mostly measured quantitatively and included interdisciplinary healthcare programmes, clinical interventions, and rehabilitation programmes focusing on pacing and breathing strategies. Compensation and insurance schemes were important funders of these interventions.

Workplace-based support was mostly measured qualitatively. Barriers to the provision of support at organisational level included lack of understanding of LC symptoms, insufficient workplace guidance, and educational gaps among managers. Individual barriers included threat of income loss, remote working and disconnection from the workplace. Facilitators for support included recognition and validation of LC and its symptoms, and eligibility for disability benefits associated with work.

**Conclusions:** RTW is an important outcome of health related absence and should be systematically recorded in studies of people with long COVID (PwLC). The heterogeneity and unpredictability of LC symptoms create challenges for supporting working age populations. Further research is crucial to better understand the specific RTW needs for PwLC and address potential barriers and facilitators to effective support. Consistent guidelines on LC’s definition, and disability status may facilitate the provision of support and the development of interventions.

**Prospero registration number:** CRD42023478126

**Article Summary:** *Strengths and limitations of this study:*

- Offers an in-depth examination of workplace and non-workplace-based support for return to work (RTW) for patients with long COVID (LC), including clinical interventions, multidisciplinary rehabilitation programmes, workplace support, and self-developed strategies.
- Comprehensive literature search of major electronic databases across disciplines and reporting as per Preferred Reporting Items for Systematic Reviews and Meta-Analyses (PRISMA) guidelines.
- The search was limited to English language, likely excluding relevant studies.
- While many studies had a low risk of bias, some studies had selection bias, unvalidated outcome measures or lacked control of confounders, limiting result accuracy.
- The focus on work-related outcomes excluded alternative support measures that might impact RTW, and the strict definition of LC omitted studies on overlapping conditions like chronic fatigue syndrome.

## Introduction

Long COVID (LC) is as a multi-system condition with symptoms persisting for 12 weeks (also known as post-COVID-19 syndrome, PSC) (1). Common symptoms include chronic fatigue, shortness of breath, difficulty concentrating, and muscle aches (2). An estimated two million people in England and Scotland (3.3% of the population) were experiencing self-reported LC as of March 2024 (3). LC was more prevalent in middle age (aged 45 to 64), females, and in those who were not working and not looking for work. Some symptoms can be quite severe and would prevent a person from trying to return to work (RTW) (4).

Supporting workers living with long-term conditions such as LC to return to work (RTW) is important because employment benefits both physical and mental health, providing financial security and fulfilling psychosocial needs (5). Unemployment is linked to higher mortality rates and poorer health outcomes (5, 6). One study found that 45% of people with LC (PwLC) had to reduce their work schedules and 22% were not working seven months after symptoms began (7).

Previous knowledge on RTW after long term sickness absence may not be easily translatable to PwLC (8). Compared to other conditions, LC symptoms often fluctuate, complicating a linear recovery process and making existing RTW measures less applicable (8, 9). LC is varyingly experienced, defined and understood, meaning individuals’ experiences of the condition, their diagnoses, treatment, and the impacts on their life may also vary substantially. This has also changed as understanding of the condition has evolved, so PwLC diagnosed in 2020 may have had difference experiences to someone diagnosed in 2025. Existing policies, especially those related to absence management, may not be adequate to meet the needs of PwLC (9). The availability and effectiveness of support for RTW for PwLC is not uniform and evidence in favour of support strategies is lacking.

This rapid review employed evidence synthesis to understand the types of RTW support and interventions, both workplace and non-workplace-based, available to PwLC. As well as barriers and facilitators to effective support for RTW, and evidence for individual strategies associated with increased RTW for PwLC.

## Methods

A review protocol was pre-published on PROSPERO (ID: CRD42023478126) and reported in line with the Preferred Reporting Items for Systematic Reviews and Meta-Analyses-Rapid Reviews (PRISMA-RR) with guidelines and adjustments made to accommodate qualitative research (10).

### Eligibility criteria

Studies describing support available for RTW for PwLC were included. LC was defined as signs and symptoms developing after acute COVID-19 infection and persisting for more than 12 weeks (1). RTW was defined as working full-time or part-time during or after LC.

Eligible studies included 1) people of working age (≥16 years old) 2) with self-reported or diagnosed LC symptoms lasting ≥12 weeks and 3) published in English. Studies of people who never had COVID-19 or with conditions overlapping with (but not including) LC (for example chronic fatigue, postural orthostatic tachycardia syndrome) were excluded.

Studies investigated 1) non-workplace based support for RTW for PwLC; 2) workplace-based support for RTW for PwLC; and 3) barriers and facilitators for those with LC RTW.

### Data sources

We searched MEDLINE, Embase, American Psychological Association (APA) PsycINFO, evidence based medicine (EBM) Reviews (including the Cochrane Central Register of Controlled Trials and Cochrane Database of Systematic Reviews), Web of Science, Google Scholar and the Health Management Information Consortium. Reference lists of relevant studies including literature reviews were hand searched for additional articles. All searches were completed in June 2024.

### Search strategy

Searches were conducted by the primary researchers (SD and HW) using synonyms for ‘long COVID’ and ‘RTW’ joined by AND operator. The search strategy for Long Covid included the terms ‘post covid’, ‘long covid’, ‘chronic covid’ and ‘post-acute covid’. For RTW, we used ‘return to work’, ‘sickness absence’, ‘employment’, ‘vocational rehabilitation’, ‘occupational rehabilitation’, ‘back to work’, ‘work participation’, ‘workability’, ‘work ability’, ‘sick leave’, ‘illness day*’, ‘disability leave’ and ‘absenteeism’. Preliminary searches were finalised in discussions with RW, MvT, DM, DF, AC and DB. The full search strategy can be found in supplementary material 1.

### Selection of studies

After removal of duplicates, studies underwent abstract and title screening against eligibility criteria by two researchers (SD and HW). Dual screening was performed for a random sample of 20% of the records to ensure consistency. The remaining records were abstract and title screened independently by one researcher. Dual full text screening of a random sample of 20% of records was conducted to obtain relevant studies. The remaining records were full text screened independently by one researcher. Differences were discussed and reconciled with input from a third author (RW) if required.

### Data extraction and quality assessment

Data from the final set of studies was extracted into an agreed proforma in Excel. The data extraction table was piloted by two researchers (SD and HW) using three studies and revised accordingly. Data extracted included study characteristics (e.g. aim, design, country), study population (e.g. demographics including age and gender, employment status, sample size, duration of LC symptoms), characteristics of interventions and comparators (e.g. type of intervention, treatment duration, patient work-related quantitative outcomes) and qualitative results (e.g. type of workplace support, providers of support if reported, barriers and facilitators to support, individual strategies for RTW).

Study quality was evaluated using the Critical Appraisal tools for use in JBI Systematic Reviews (11). The checklists include subscale items that were evaluated with ‘yes’, ‘no’ or ‘unclear’. Risk of bias was described as low, moderate, or high quality based on the tool. Any disagreements were resolved by discussion or by involving another author (RW).

### Data synthesis

A narrative synthesis was performed to present both quantitative and qualitative results. Due to the diverse nature of intervention descriptions and outcome measures, conducting a meta-analysis was unfeasible. This approach allowed for a structured comparison of workplace and non-workplace RTW support while integrating both quantitative work-related outcomes and qualitative insights. Summary tables facilitated exploration of clinical contexts, intervention components, and RTW impacts, while thematic analysis of qualitative studies provided a deeper understanding of organisational and individual barriers and facilitators to RTW for PwLC.

### Patient and Public Involvement

Scoping questions and research priorities were co-developed with a local LC patient support group and a team member with lived experience (DF).

## Results

### Search results

We identified 1,036 records after deduplication: 983 from the database search and 53 from citation screening. Following title and abstract screening, 299 articles underwent full-text screening and 25 met eligibility criteria for inclusion (Figure 1).

**Figure 1:**
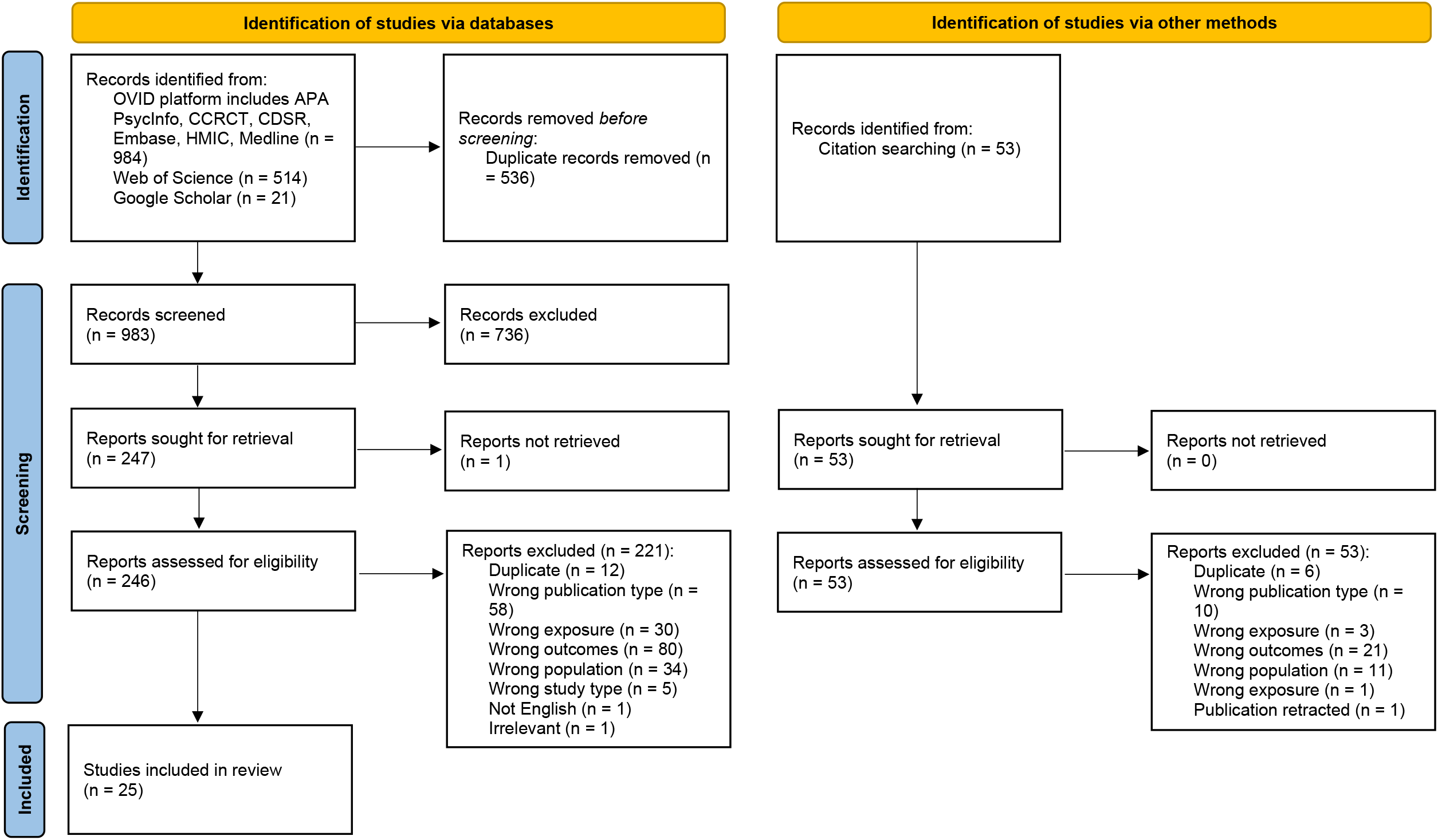
PRISMA flow diagram

### Study characteristics

Characteristics of the included studies are presented in supplementary material 2. All studies were published between 2021 and 2024. Of the 25 studies, two were cross-sectional (12, 13), five were prospective cohort studies (14-18), three were retrospective cohort studies (19-21), one was a quasi-experimental study because it assessed the effects of an intervention on participants without randomisation (22), two were case studies (23, 24), ten were qualitative studies (8, 25-33), and two were randomised controlled trials (RCTs) (34, 35). All studies were published between 2021-2023 and predominantly conducted in Europe (n=17) and North America (n=6), only one in Asia and one was multi-national.

The studies included 2,964 participants, ranging from 50% and 100% female and aged 18 to >75 years old. Where ethnicity was reported, the majority included mostly white ethnic groups (ranging from 68% to 87% white ethnic groups in four studies) (25, 29, 30, 32).

### Quality assessment

The assessed studies were heterogenous in design and applicability to the general population. Fifteen of the studies were identified as low risk of bias based on description of methodology and reliability of reportable outcomes, although a number of studies categorised as low risk were case studies or quasi-experimental so liable to selection bias (supplementary material 2).

The two case studies were assessed as low risk of bias within their own study design (23, 24). Both studies clearly described the participant demographics and clinical history, and the rehabilitation methods were well described. The two RCTs had a medium risk of bias due to selection and allocation issues, with non-concealed allocation in one study and unclear blinding in both and a lack of baseline similarity in group characteristics at the start of both studies (34, 35). Among the cohort studies, four were low risk (16, 17, 19, 21), three medium (14, 15, 18), and one high risk (20). All studies had one or more limitations, including unclear LC ascertainment, lack of confounder control, unreported or unclear measurement tool validity, and unaddressed incomplete follow-up. The quasi-experimental cohort study was low risk but faced limited compatibility due to the absence of a matched control group, though they used reliable pre- and post-intervention measurements with minimal participant loss to follow-up (22). The two cross-sectional studies were considered low risk but one did not explicitly mention specific strategies to deal with confounding factors (13), and the other study had unclear measurement tool validity (12). Among the qualitative studies, seven were low risk (8, 25, 26, 28, 29, 32, 33), two medium (30, 31), and one high risk (27). Most studies showed congruity between the research question, methodology, and philosophical perspective, except for Kohn et al (27), which did not adequately represent participant voices. Only two studies considered the researchers’ potential influence on the research (8, 29).

### Workplace support for RTW with LC, and barriers and facilitators to support

Thirteen studies reported on workplace-based support for RTW with LC (8, 12, 13, 23, 25-33) (supplementary material 3). The findings were primarily based on qualitative data collected through methods such as focus groups, interviews, case studies, or open-ended survey questions. The studies gathered data from the perspectives of PwLC, focusing on their lived experiences and expectations, including suggestions for support. The suggestions were made for general conditions of LC without specifying symptoms. One study used a cross-sectional design to provide quantitative survey data on how many PwLC found workplace support to be helpful or not (13).

Commonly described workplace support measures included adjusting work arrangements, such as reducing or offering flexible work hours, allowing breaks, enabling working from home (WFH) or teleworking, adjusting job content or roles, and sharing workloads. Emotional support was reported, such as good communication, regular check-ins, and demonstrating understanding toward employees’ needs (8, 25, 26, 30). Financial support was also reported, such as where a condition was acquired at work, through compensation claims, typically provided by employers or their insurance carriers, as well as government financial support (29, 33). Some participants proposed vocational integration programmes as a practical support to help individuals reintegrate into the workplace after a period of absence (28).

We categorised ‘organisational barriers’ as barriers to the provision of workplace support for RTW placed on the PwLC by the organisation, rather than by the individual. Organisational barriers to the provision of support included the lack of recognition, understanding or knowledge about LC and related symptoms (8, 25-27, 29, 30, 32, 33). This resulted in reports of workplace stigma associated with LC (25). Additionally, flexibility was sometimes not provided due to workplace constraints (12).

Workplace culture emerged as a major concern, with organisational barriers stemming from management and colleague perceptions of participants’ ability to work. For example, a lack of understanding about the participants’ illness which could lead to friction and hostility (33), and insensitive attitudes from coworkers (31). Participants described a toxic environment where colleagues showed little interest in understanding the LC condition, suspicion about avoiding work, and alienation (8, 25, 31). Participants also described pressure from management to ignore symptoms (8) and questioning the legitimacy of COVID-19 health consequences (28).

We categorised ‘individual barriers’ as emotional and financial stressors that limit the provision of workplace support for RTW in PwLC. Some participants faced individual barriers such as strain, guilt, anxiety, negative self-perception regarding their work abilities, and identity crises (8, 25, 27, 28, 31, 33). Work adjustments, such as reduced hours or medical leave, were associated with income loss, posing a significant concern and, thus, and individual barrier to obtaining such support (8, 27, 28). When WFH, some individuals were concerned about limited or no contact with colleagues and the difficulties in establishing new connections (28).

Organisational facilitators to the provision of support included recognising LC as a legitimate condition, fostering empathy among colleagues and management, and providing access to disability benefits (8, 25, 27, 31). Organisational policies addressing the needs of PwLC, education initiatives to equip staff with knowledge on how to support them, and workplace support groups to promote advocacy within organisations were also highlighted (25, 31). Additionally, occupational health services recognising individual needs and capabilities was also considered a facilitator to gaining support (31).

Straßburger et al (13) surveyed organisational support for RTW with LC, finding that structural changes - specifically reduced working hours, workplace adjustments, and task modifications - were considered helpful to facilitate successful reintegration into work-life, while occupational reintegration plans, health courses, general consultation, and job coaching were less helpful.

Tan and Koh (23) reported a case study on a participant who developed their own rehabilitation programme due to the lack of occupational health services provided by their workplace. Other individual strategies included finding a good balance between activity and rest (26), adopting a pragmatic approach to symptom management by not hiding the symptoms (25), communication strategies to manage expectations (31), knowing one’s employment rights (31), and redefining occupational identity to find self-worth (8).

### Non-workplace-based support for return to work with long COVID

Twelve studies investigated non-workplace-based support for PwLC to RTW, including clinical interventions, rehabilitation programmes and multidisciplinary programmes (14-22, 24, 34, 35). Most of the intervention studies used longitudinal or experimental designs, except for Wagner et al (24), a single participant case report. Generally, interventions were effective, though Müller et al (17) and Kerling et al (35) found no significant differences between groups. The Work Ability Index (WAI) was the most frequently used outcome measure. The results are summarised in supplementary material 4.

Most of the intervention programmes supported all PwLC with a variety of symptoms, although one targeted neurocognitive impairments (i.e. brain fog) (20) and two targeted fatigue (16, 24).

Derksen et al (34) evaluated an RCT of an interdisciplinary health care facilitation programme, including medical internet support from personal pilots and digital interventions. The primary focus of the intervention was to address a broader set of clinical goals, including symptom reduction and enhanced social participation. RTW outcomes were part of the evaluation, specifically through the assessment of work ability. They divided participants into three groups: an intervention group (IG), an active control group (ACG), and a comparison group (CompG), which received standard care without specific treatment. Both the IG and ACG received professional personal pilot support and digital interventions via a medical internet aid, while the IG also underwent a 3-day diagnostic assessment at a specialised neurological rehabilitation clinic. Outcomes were assessed at five points: initial screening (T1), 4 weeks (T2), 6 weeks of intervention (T3, IG only), post-intervention (10-12 weeks, T4), and 6 weeks post-intervention (T5). The WAI scores improved for both IG and ACG at T4 and T5 compared to T2, with CompG showing slight improvement at T4. Both IG and ACG demonstrated higher work ability than CompG, although no significant difference was observed between IG and ACG over time.

LeGoff et al (20) carried out a neurocognitive screening evaluation (NCSE) to assess symptoms and disabilities associated with post–COVID-19 condition (PCC) and facilitating employee recovery and RTW. The NCSE functioned as both a diagnostic tool and a therapeutic intervention, providing objective feedback that may have reassured employees about their recovery potential. Its aim was to help streamline treatment recommendations and RTW decisions, leading to faster recovery timelines. Following the NCSE, the loss of workdays significantly decreased compared to pre-referral levels (20). The NCSE, adapted from a previous study on post-concussion cases (36), served both as an assessment and an intervention by providing objective feedback to patients. Frisk et al (22) reported on a clinical intervention that included a phase for integrating rehabilitation changes into daily life, leading to a 20% reduction in sick leave among employed participants after three months. Tanguay et al (15) reported that a telerehabilitation programme utilising pacing strategies and occupational therapy reduced the number of participants on medical leave by week 8 and increased the number of participants returning to full-time work.

Altmann et al. (2023) (14) examined a rehabilitation programme that incorporated multimodal respiratory therapy, endurance and resistance muscle training, psychological support, and educational interventions, tailored for COVID-19 and LC patients. The study found that at discharge, a higher proportion of LC participants (infected around 10 months prior) were rated as fit for work immediately compared to COVID-19 participants (infected 2 months prior). However, more COVID-19 participants were expected to RTW within 6 months. This suggests that LC participants can eventually reach a fitness level comparable to or better than recent COVID-19 participants, but there is a longer recovery period observed in some LC participants. Altmann et al also reported that for LC participants, difficulties in RTW were primarily due to neurocognitive pathology rather than pulmonary impairment (14).

Brehon et al (19) focused on workers who filed LC compensation claims due to workplace COVID-19 exposure. Their multidisciplinary programme, which included various therapies and psychoeducation, effectively improved RTW rates at discharge. Of those returning, 7% resumed regular duties, while 93% returned to modified duties (19). In addition, those who reported having modified duties available at their workplace were 3.38 times more likely to RTW upon programme discharge compared to those without such options (19).

Ghali et al (21) followed patients with post-COVID-19 syndrome (PCS) adhering to pacing strategies for recovery. Patients who adhered more strictly to pacing strategies (i.e. high engagement in pacing subscale scores) experienced higher recovery rates, defined as the ability to RTW.

Kerling et al (35) evaluated a 3-month home exercise plan with moderate and intense activities using the WAI. The intervention showed no change in work ability from baseline.

Kvale et al (18) compared a micro-choice-based rehabilitation across three groups: low back pain, LC, and type 2 diabetes. The programme involved three phases: (1) preparing for change, (2) a concentrated 3-4 day intervention, and (3) integrating changes into daily life. Patients were taught and practiced how to monitor and target seemingly insignificant everyday micro-choices to break behavioural patterns that reduced functionality or contributed to health problems. Functioning improved for low back pain and LC patients, but not for diabetes patients, as measured by the Work and Social Adjustment Scale.

Schmid et al (16) reported on a 14-day multimodal integrative inpatient programme, offering personalised therapies including vitamins, hydrotherapy, thermotherapy, and mind-body medicine. Self-reported work ability significantly increased after treatment and six months later.

## Discussion

This rapid review sought to address the key questions: what support, if any, is being provided that assists PwLC RTW and what barriers and facilitators exist to accessing this support.

### Interventions that improve work ability of people with long COVID

Our findings have identified several interventions and rehabilitation programmes supporting PwLC in RTW. These include an interdisciplinary healthcare facilitation programme with digital technology support, clinical interventions that integrate improvements into daily living, multidisciplinary programmes combining therapy and medical interventions, and rehabilitation programmes offering guidance on pacing and breathing strategies (14-16, 19, 22, 34).

While most interventions positively influenced RTW outcomes, Kerling et al (35) noted no significant WAI improvement following a home exercise plan, highlighting the limitations of low-intensity interventions, that lack professional oversight. Motivation and adherence could play a significant role in the success of any intervention, particularly unmonitored ones (37). Low-intensity home exercises can seem monotonous over time, potentially leading to decreased engagement (38). Furthermore, individuals recovering from illnesses like LC may encounter fatigue, post-exertional malaise or mood disturbances, which may impact participation in physical exercise (37-39).

Derksen et al’s study (34) found that both the IG and ACG groups showed improved WAI scores compared to the standard care group, but the tailored diagnostic assessment in the IG offered no additional long-term benefits. While the authors highlighted positive feedback from the IG participants regarding the clinical assessment; they noted that accessing and completing specialist and rehabilitative care tailored to individual needs could be a long-term process, suggesting that the shorter follow-up period in the study may not have been sufficient to capture the intervention’s full effects. Similarly, Brehon et al (19) reported no immediate improvements in work ability following occupational rehabilitation for PwLC. Over one year, however, significant health gains and work reintegration outcomes were observed, indicating that longer assessment periods are needed to evaluate the full impact of some tailored rehabilitation programmes. Furthermore, Brehon et al (19) showed that the presence of modified duties in the workplace at admission to LC rehabilitation results in better RTW outcomes, suggesting that workplace adjustments are crucial alongside rehabilitation efforts. The need for longer periods to assess RTW may reflect the fluctuating nature of LC and RTW should be included in study outcomes at inception to develop evidence for effective interventions.

Globally, definitions, descriptions and solutions for LC vary (40). The 2024 ‘National Academies of Sciences, Engineering, and Medicine Long COVID Definition’ aimed to address these issues by providing a comprehensive framework that includes a wide range of symptoms and conditions (41). Definitions prior to this time were less established, impacting comparison and applicability of studies done in this period. Ambiguity around definitions of LC have influenced policy, legal and regulatory responses, and may have altered individuals’ workplace rights. In the UK, for example, LC is classified as a disability under the Equality Act only if it has ‘substantial impact on a person’s life’ or is expected to last 12 months or more (42). As a result, employers may not be legally obligated to provide reasonable adjustments for individuals with LC. This may lead to different treatment for PwLC in attempted RTW and contribute to differing outcomes of successful RTW in studies.

The majority of the included intervention studies in this review were funded by compensation or insurance providers in high-income countries, e.g. Germany, Canada and the US (14, 15, 17, 19, 20, 34). In these countries workplace acquired COVID-19 infection and LC are treated as occupational diseases or work-related accidents and hence compensable (43-46). This recognition as a compensable condition may provide a financial incentive for insurance providers to invest in research and intervention development. The involvement of insurance providers in funding LC interventions could drive significant advancements in understanding and managing the condition. Conversely, it may also lead to a focus on cost-effective solutions that prioritise short-term work-related outcomes over comprehensive long-term care. Additionally, these schemes require contributions from workers, employers, and sometimes the government, making them inaccessible to those who cannot afford to pay into them. If someone has lost their job, they may not benefit from these schemes. Furthermore, these schemes are not universal and may not be available in all countries. Moreover, many of these interventions were delivered within healthcare settings. Persons with disabilities can face barriers to accessing healthcare interventions due to financial constraints, lack of health insurance, geographical limitations or demanding work schedules, making it difficult for them to attend regular appointments, among many other reasons (47). Therefore, they are unlikely to help everyone with LC to RTW, indicating a need for complementary provisions.

### Barriers and facilitators to return to work support

Workplace flexibility and adaptability, understanding, compassion and communication were identified as important in facilitating RTW for PwLC. The included qualitative studies described support for RTW such as reducing work hours, flexible scheduling, WFH, altering job content or roles, good communication, and emotional and financial support (12, 25, 26, 28, 29). These suggestions align with current UK guidance for LC which recommend that employers maintain communication and provide flexible work arrangements, such as phased returns and task adjustments, with a focus on creating a supportive workplace culture to accommodate fluctuating symptoms (48, 49). Good working relationships, particularly between line managers and individuals, have been highlighted as crucial in enabling RTW for individuals with health problems (50, 51). UK Government financial support includes eligibility for benefits like Statutory Sick Pay and Access to Work (52). These suggestions also align with those recommended for supporting workers with long-term conditions experiencing similar symptoms such as chronic fatigue syndrome (53, 54).

Included studies investigating the lived experiences of LC workers consistently reported that lack of understanding about LC symptoms, insufficient guidance or knowledge, lack of education for managers and workplaces, and workplace stigma presented major barriers to the provision of good quality RTW support (25-27, 29). These findings support existing research on experiences of LC (55). Damant et al (56) developed a PCC-related stigma measurement and found that stigma scores were significantly associated with the number, intensity, and burden of symptoms. Furthermore, workplace adjustments may come with unintended consequences such as loss of income if working hours are reduced (27, 28), or feelings of disconnectedness when WFH (28). Straβburger et al (13) found that organisation structural changes in the physical or logistical aspects of a workplace, such as modifying the workplace environment, adjusting working hours, or changing the nature of employee tasks, were found to be the more helpful than other measures like job coaching or health courses, that are more supportive or educational in nature. However, the study did not investigate the reasons for these results. Conversely, other studies have shown that job coaching and psychoeducational interventions can effectively help manage chronic conditions by improving perceptions of working capacity, self-efficacy, and fatigue (57-59).

The heterogeneity of LC symptoms and their unpredictable nature make it challenging to develop universally effective support for RTW (9). Findings from this review demonstrate that some individuals develop their own individual coping strategies, such as balancing activity and rest, reorganising home responsibilities, and openly managing symptoms. Varying and non-specific symptoms in other conditions have been effectively managed through education and strategies for self-management and mindfulness interventions (60-62). This review suggests a need for the development of and evaluation of targeted self-management programmes to facilitate RTW for those affected by LC. This should be balanced with the responsibility of employers to make workplace accommodations.

### Implications for research and policy

Symptoms of LC are not universal to all patients, multi-system, not readily visible, and varied among individuals (for example a person may experience predominantly neurological or respiratory symptoms) and over time. The unpredictability of day-to-day symptoms and differences in recovery trajectories makes a ‘one size fits all’ approach to RTW workplace support and interventions challenging. Indeed, this is true for workplace support for other chronic or recurring conditions (63).

Our findings demonstrate that a successful RTW for PwLC requires a multidisciplinary approach involving managers, human resources, health professionals, disability support and compensation systems and a critical role for occupational health in coordinating these efforts (64). A tailored, long-term, and flexible RTW plan is essential to accommodate the complex nature of the condition. Unfortunately, only half of the UK population has access to occupational health services, and work adjustments are not universally available or always feasible (65). For example, workers in small and medium-sized enterprises (SMEs) generally have less access to occupational health services (66). SMEs are also more reluctant to invest the time and resources into making the necessary adjustments (66) which could lead to more job insecurity and pay differences. As a result, PwLC may experience significant barriers to necessary support to RTW and maintain their employment.

The insights gained from this review could inform policy development to help workers to return to their roles, either partially or fully, more effectively. The studies collectively affirm the effectiveness of tailored, multidisciplinary strategies in improving RTW outcomes for individuals with LC compared to less integrative approaches. The research highlights the importance of workplace adjustments, such as modified duties, and calls for scalable, evidence-based solutions that combine individualised interventions, supportive workplace environments, and ongoing evaluation. These findings may also apply to managing other chronic conditions with similar symptomology, such as chronic fatigue syndrome. There may also be valuable lessons to be learned from these conditions too, which could inform strategies for supporting PwLC.

This review focussed on both non-workplace and workplace-based support for RTW with LC. We found that non-workplace-based support tended to be reported by quantitative clinical studies with effectiveness considered. While support provided by the workplace was mostly reported by qualitative studies where evaluation of effectiveness was more difficult. Similar to other long-term conditions, workplace support is much needed for returning to or remaining at work (5, 27). Evaluating complex interventions outside of clinical settings remains a challenge for public health researchers (67).

### Strength and limitations

This review provides a comprehensive report of workplace- and non-workplace-based support for RTW with LC. The data extracted covers clinical interventions, multidisciplinary rehabilitation programmes, workplace support and self-developed individual support programmes. It benefits from a comprehensive literature search across major electronic databases and adheres to the rigorous reporting standards of the PRISMA guidelines. However, the search was limited to English-language studies, which may have excluded some relevant research. Additionally, the focus on work-related outcomes means that we did not include alternative support for LC patients where there may have been an impact on RTW.

While many studies were assessed as low risk of bias, several still had inherent limitations, particularly case studies and quasi-experimental designs, which are prone to selection bias. Many of the non-workplace based studies did not sufficiently address potential confounders, limiting the ability to attribute work outcomes solely to the intervention or other factors. For example, differences in baseline health status, comorbidities or vaccination status that may have affected recovery trajectories (14-16, 18-20). Also, some studies employed outcome measures without clearly indicating whether they used validated tools to assess LC-related RTW or work ability status (16, 20, 21). This inconsistency raises concerns about the accuracy and interpretation of findings.

The strict definition of LC precluded other studies that focused on conditions overlapping with LC symptoms, e.g. chronic fatigue or neurocognitive conditions. Learning on barriers and facilitators to RTW in these conditions may also be relevant to those with LC. Additionally, LC definitions and access to support have changed considerably since it was recognised in mid-2020. This review did not consider these different time periods, which could impact the findings.

## Conclusion

Non-workplace-based interventions, such as interdisciplinary healthcare programmes and rehabilitation focusing on pacing and breathing strategies, were often funded by compensation and insurance schemes. Workplace-based support, primarily explored qualitatively, revealed significant organisational barriers, including a lack of understanding of LC symptoms, inadequate workplace guidance, and limited manager education. Key facilitators included acknowledging and validating LC symptoms and eligibility for disability benefits.

The heterogeneity and unpredictability of LC symptoms pose significant challenges for supporting working age individuals. Further research is needed to uncover the specific RTW needs of people with LC, while addressing barriers and leveraging facilitators. Establishing consistent guidelines on LC’s definition, compensation, and disability status could enhance support efforts and guide intervention development. Acknowledging RTW as a key health outcome and consistently documenting it in research on PwLC is crucial for driving progress in this field.

## Acknowledgements

We thank members of the Long COVID Patient Support Group Greater Manchester for their contribution to the overall study design, research methods and the strategies for dissemination. Their input was valuable in ensuring that the findings would reach and resonate with diverse audiences, including those directly affected by long COVID.

## Author Contributions

SD and HW contributed to conception and design of the work, data collection, data analysis and original draft preparation. RW contributed to supervision, conception and design of the work, data collection, draft review and editing. MvT contributed to supervision, conception and design of the work, draft review and editing. DF, DM, AC and DB contributed to conception and design of the work, draft review and editing.

## Patient consent for publication

Not required.

## Data availability statement

Data are available on reasonable request. All data relevant to the study are included in the article uploaded as supplementary information or deposited on Open Science Framework, DOI: https://osf.io/rvzd7/?view_only=cc7142b7f5234824b19391d193ab53e3.

## References

1. National Institute for Health and Excellence. Long-term effects of coronavirus (long COVID): What is it? [online]. 2022. https://cks.nice.org.uk/topics/long-term-effects-of-coronavirus-long-covid/background-information/definition/#:∼:text=It%20includes%20both%20ongoing%20symptomatic,syndrome%2C%20and%20permanent%20organ%20damage (accessed 30 April 2024).

2. National Institute for Health and Excellence. Long-term effects of coronavirus (long COVID): How common is it? [online]. 2022. https://cks.nice.org.uk/topics/long-term-effects-of-coronavirus-long-covid/background-information/prevalence/ (accessed 13 May 2024).

3. Office for National Statistics. Self-reported coronavirus (COVID-19) infections and associated symptoms, England and Scotland: November 2023 to March 2024 [online]. 2024. https://www.ons.gov.uk/peoplepopulationandcommunity/healthandsocialcare/conditionsanddiseases/articles/selfreportedcoronaviruscovid19infectionsandassociatedsymptomsenglandandscotland/november2023tomarch2024 (accessed 13 May 2024).

4. Industrial Injuries Advisory Council. Industrial Injuries Advisory Council independent report: Covid-19 and occupational impacts [online]. 2022. https://www.gov.uk/government/publications/covid-19-and-occupational-impacts (accessed 14 May 2024).

5. Waddell G, Burton KA. Is work good for your health and well-being? An independent review [online]. 2006. https://www.gov.uk/government/publications/is-work-good-for-your-health-and-well-being (accessed 13 May 2024).

6. Barnes MC, Gunnell D, Davies R, Hawton K, Kapur N, Potokar J, et al. Understanding vulnerability to self-harm in times of economic hardship and austerity: a qualitative study. BMJ Open. 2016;6(2):e010131.

7. Davis HE, Assaf GS, McCorkell L, Wei H, Low RJ, Re’em Y, et al. Characterizing long COVID in an international cohort: 7 months of symptoms and their impact. EClinicalMedicine. 2021;38:101019.

8. Nielsen K, Yarker J. “It’s a rollercoaster”: the recovery and return to work experiences of workers with long COVID. Work & Stress. 2024;38(2):202–30.

9. Lunt J, Hemming S, Elander J, Baraniak A, Burton K, Ellington D. Experiences of workers with post-COVID-19 symptoms can signpost suitable workplace accommodations. International Journal of Workplace Health Management. 2022;15(3):359–74.

10. Stevens A, Garritty C, Hersi M, Moher D. Developing PRISMA-RR, a reporting guideline for rapid reviews of primary studies (Protocol). Equator Network. 2018.

11. Aromataris E, Munn Z. JBI Manual for Evidence Synthesis: JBI [online]. 2020. https://synthesismanual.jbi.global. z10.46658/JBIMES-24-01 (accessed 13 May 2024).

12. Delgado-Alonso C, Cuevas C, Oliver-Mas S, Díez-Cirarda M, Delgado-Álvarez A, Gil-Moreno MJ, et al. Fatigue and Cognitive Dysfunction Are Associated with Occupational Status in Post-COVID Syndrome. International Journal of Environmental Research and Public Health [Internet]. 2022; 19(20).

13. Straßburger C, Hieber D, Karthan M, Jüster M, Schobel J. Return to work after Post-COVID: describing affected employees’ perceptions of personal resources, organizational offerings and care pathways. Front Public Health. 2023;11:1282507.

14. Altmann CH, Zvonova E, Richter L, Schüller PO. Pulmonary recovery directly after COVID-19 and in Long-COVID. Respiratory Physiology & Neurobiology. 2023;315:104112.

15. Tanguay P, Gaboury I, Daigle F, Bhéreur A, Dubois O, Lagueux É, et al. Post-exertional malaise may persist in Long COVID despite learning STOP-REST-PACE. Fatigue: Biomedicine, Health & Behavior. 2023;11(2-4):113–28.

16. Schmid S, Uecker C, Fröhlich A, Langhorst J. Effects of an integrative multimodal inpatient program on fatigue and work ability in patients with Post-COVID Syndrome-a prospective observational study. Eur Arch Psychiatry Clin Neurosci. 2024.

17. Müller K, Poppele I, Ottiger M, Zwingmann K, Berger I, Thomas A, et al. Impact of Rehabilitation on Physical and Neuropsychological Health of Patients Who Acquired COVID-19 in the Workplace. International Journal of Environmental Research and Public Health [Internet]. 2023; 20(2).

18. Kvale G, Søfteland E, Jürgensen M, Wilhelmsen-Langeland A, Haugstvedt A, Hystad SW, et al. First trans-diagnostic experiences with a novel micro-choice based concentrated group rehabilitation for patients with low back pain, long COVID, and type 2 diabetes: a pilot study. BMC Med. 2024;22(1):12.

19. Brehon K, Niemeläinen R, Hall M, Bostick GP, Brown CA, Wieler M, et al. Return-to-Work Following Occupational Rehabilitation for Long COVID: Descriptive Cohort Study. JMIR Rehabil Assist Technol. 2022;9(3):e39883.

20. LeGoff DB, Lazarovic J, Kofeldt M, Peters A. Neurocognitive and Symptom Validity Testing for Post–COVID-19 Condition in a Workers Compensation Context. Journal of Occupational and Environmental Medicine. 2023;65(10).

21. Ghali A, Lacombe V, Ravaiau C, Delattre E, Ghali M, Urbanski G, et al. The relevance of pacing strategies in managing symptoms of post-COVID-19 syndrome. Journal of Translational Medicine. 2023;21(1):375.

22. Frisk B, Jürgensen M, Espehaug B, Njøten KL, Søfteland E, Aarli BB, et al. A safe and effective micro-choice based rehabilitation for patients with long COVID: results from a quasi-experimental study. Sci Rep. 2023;13(1):9423.

23. Tan KWA, Koh D. Long COVID—Challenges in diagnosis and managing return-to-work. Journal of Occupational Health. 2023;65(1):e12401.

24. Wagner B, Steiner M, Markovic L, Crevenna R. Successful application of pulsed electromagnetic fields in a patient with post-COVID-19 fatigue: a case report. Wiener Medizinische Wochenschrift. 2022;172(9):227–32.

25. Chasco EE, Dukes K, Jones D, Comellas AP, Hoffman RM, Garg A. Brain Fog and Fatigue following COVID-19 Infection: An Exploratory Study of Patient Experiences of Long COVID. International Journal of Environmental Research and Public Health [Internet]. 2022; 19(23).

26. Gyllensten K, Holm A, Sandén H. Workplace factors that promote and hinder work ability and return to work among individuals with long-term effects of COVID-19: A qualitative study. Work. 2023;75:1101–12.

27. Kohn L, Dauvrin M, Detollenaere J, Primus-de Jong C, Maertens de Noordhout C, Castanares-Zapatero D, et al. Long COVID and return to work: a qualitative study. Occup Med (Lond). 2024;74(1):29–36.

28. Schmachtenberg T, Müller F, Kranz J, Dragaqina A, Wegener G, Königs G, et al. How do long COVID patients perceive their current life situation and occupational perspective? Results of a qualitative interview study in Germany. Frontiers in Public Health. 2023;11.

29. Stelson EA, Dash D, McCorkell L, Wilson C, Assaf G, Re’em Y, et al. Return-to-work with long COVID: An Episodic Disability and Total Worker Health® analysis. Social Science & Medicine. 2023;338:116336.

30. Folk AL. Exploring the experiences of academic library employees with long COVID in the United States and Canada. The Journal of Academic Librarianship. 2023;49(6):102790.

31. Lunt J, Hemming S, Elander J, Burton K, Hanney B. Sustaining work ability amongst female professional workers with long COVID. Occup Med (Lond). 2024;74(1):104–12.

32. McNabb KC, Bergman AJ, Smith-Wright R, Seltzer J, Slone SE, Tomiwa T, et al. “It was almost like it’s set up for people to fail” A qualitative analysis of experiences and unmet supportive needs of people with Long COVID. BMC Public Health. 2023;23(1):2131.

33. Miller A, Song N, Sivan M, Chowdhury R, Burke MR. Identifying the needs of people with long COVID: a qualitative study in the UK. BMJ Open. 2024;14(6):e082728.

34. Derksen C, Rinn R, Gao L, Dahmen A, Cordes C, Kolb C, et al. Longitudinal Evaluation of an Integrated Post–COVID-19/Long COVID Management Program Consisting of Digital Interventions and Personal Support: Randomized Controlled Trial. J Med Internet Res. 2023;25:e49342.

35. Kerling A, Beyer S, Dirks M, Scharbau M, Hennemann AK, Dopfer-Jablonka A, et al. Effects of a randomized-controlled and online-supported physical activity intervention on exercise capacity, fatigue and health related quality of life in patients with post-COVID-19 syndrome. BMC Sports Sci Med Rehabil. 2024;16(1):33.

36. LeGoff DB, Wright R, Lazarovic J, Kofeldt M, Peters A. Improving Outcomes for Work-Related Concussions: A Mental Health Screening and Brief Therapy Model. Journal of Occupational and Environmental Medicine. 2021;63(10).

37. Bachmann C, Oesch P, Bachmann S. Recommendations for Improving Adherence to Home-Based Exercise: A Systematic Review. Physikalische Medizin, Rehabilitationsmedizin, Kurortmedizin. 2017;28.

38. Jurkiewicz MT, Marzolini S, Oh P. Adherence to a Home-Based Exercise Program for Individuals After Stroke. Topics in Stroke Rehabilitation. 2011;18(3):277–84.

39. Gloeckl R, Zwick RH, Fürlinger U, Schneeberger T, Leitl D, Jarosch I, et al. Practical Recommendations for Exercise Training in Patients with Long COVID with or without Post-exertional Malaise: A Best Practice Proposal. Sports Medicine - Open. 2024;10(1):47.

40. Munblit D, O’Hara ME, Akrami A, Perego E, Olliaro P, Needham DM. Long COVID: aiming for a consensus. Lancet Respir Med. 2022;10(7):632–4.

41. National Academies of Sciences E, Medicine, Health, Medicine D, Board on Global H, Board on Health Sciences P, et al. In: Goldowitz I, Worku T, Brown L, Fineberg HV, editors. A Long COVID Definition: A Chronic, Systemic Disease State with Profound Consequences. Washington (DC): National Academies Press (US).

42. Anderson E, Hunt K, Wild C, Nettleton S, Ziebland S, MacLean A. Episodic disability and adjustments for work: the ‘rehabilitative work’ of returning to employment with Long Covid. Disability & Society. 2024:1–23.

43. Sandal A, Yildiz AN. COVID-19 as a Recognized Work-Related Disease: The Current Situation Worldwide. Safety and Health at Work. 2021;12(1):136–8.

44. U.S. Department of Labor. Claims under the Federal Employees’ Compensation Act due to COVID-19 [online]. 2022. https://www.dol.gov/agencies/owcp/FECA/InfoFECACoverageCoronavirus (accessed 8 January 2025).

45. Nienhaus A. COVID-19 among Health Workers in Germany-An Update. Int J Environ Res Public Health. 2021;18(17).

46. Smith PM, Liao Q, Shahidi F, Biswas A, Robson LS, Landsman V, et al. Variation in occupational exposure risk for COVID-19 workers’ compensation claims across pandemic waves in Ontario. Occupational and Environmental Medicine. 2024;81(4):171.

47. Gréaux M, Moro MF, Kamenov K, Russell AM, Barrett D, Cieza A. Health equity for persons with disabilities: a global scoping review on barriers and interventions in healthcare services. International Journal for Equity in Health. 2023;22(1):236.

48. CIPD. Working with long COVID: Guidance for people professionals [online]. 2024. https://www.cipd.org/en/knowledge/guides/long-covid-guides/ (accessed 17 January 2025).

49. Society of Occupational Medicine. LONG COVID: A Manager’s guide [online]. 2024. https://www.som.org.uk/sites/som.org.uk/files/Long_COVID_and_Return_to_Work_What_Works.pdf (accessed 17 January 2025).

50. Nielsen K, Yarker J. What can I do for you? Line managers’ behaviors to support return to work for workers with common mental disorders. Journal of Managerial Psychology. 2022;38.

51. Cohen D, Allen J, Rhydderch M, Aylward M. The return to work discussion: a qualitative study of the line manager conversation about return to work and the development of an educational programme. J Rehabil Med. 2012;44(8):677–83.

52. Department of Health and Social Care. Guidance: Find help and support if you have long COVID [online]. 2024. https://www.gov.uk/guidance/find-help-and-support-if-you-have-long-covid (accessed 17 January 2025).

53. Ali S, Chalder T, Madan I. Evaluating Interactive Fatigue Management Workshops for Occupational Health Professionals in the United Kingdom. Safety and Health at Work. 2014;5(4):191–7.

54. Stevelink SAM, Mark KM, Fear NT, Hotopf M, Chalder T. Chronic fatigue syndrome and occupational status: a retrospective longitudinal study. Occupational Medicine. 2022;72(3):177–83.

55. Michelen M, Manoharan L, Elkheir N, Cheng V, Dagens A, Hastie C, et al. Characterising long COVID: a living systematic review. BMJ Global Health. 2021;6(9):e005427.

56. Damant RW, Rourke L, Cui Y, Lam GY, Smith MP, Fuhr DP, et al. Reliability and validity of the post COVID-19 condition stigma questionnaire: a prospective cohort study. eClinicalMedicine. 2023;55:101755.

57. Nazarov S, Manuwald U, Leonardi M, Silvaggi F, Foucaud J, Lamore K, et al. Chronic Diseases and Employment: Which Interventions Support the Maintenance of Work and Return to Work among Workers with Chronic Illnesses? A Systematic Review. International Journal of Environmental Research and Public Health [Internet]. 2019; 16(10).

58. Varekamp I, Verbeek JH, de Boer A, van Dijk FJH. Effect of job maintenance training program for employees with chronic disease — a randomized controlled trial on self-efficacy, job satisfaction, and fatigue. Scandinavian Journal of Work, Environment & Health. 2011;37(4):288–97.

59. Varekamp I, Krol B, van Dijk FJH. Empowering employees with chronic diseases: process evaluation of an intervention aimed at job retention. International Archives of Occupational and Environmental Health. 2011;84(1):35–43.

60. Acabchuk RL, Brisson JM, Park CL, Babbott-Bryan N, Parmelee OA, Johnson BT. Therapeutic Effects of Meditation, Yoga, and Mindfulness-Based Interventions for Chronic Symptoms of Mild Traumatic Brain Injury: A Systematic Review and Meta-Analysis. Appl Psychol Health Well Being. 2021;13(1):34–62.

61. Bernstein LJ, McCreath GA, Nyhof-Young J, Dissanayake D, Rich JB. A brief psychoeducational intervention improves memory contentment in breast cancer survivors with cognitive concerns: results of a single-arm prospective study. Support Care Cancer. 2018;26(8):2851–9.

62. Gok Metin Z, Karadas C, Izgu N, Ozdemir L, Demirci U. Effects of progressive muscle relaxation and mindfulness meditation on fatigue, coping styles, and quality of life in early breast cancer patients: An assessor blinded, three-arm, randomized controlled trial. Eur J Oncol Nurs. 2019;42:116–25.

63. Pransky GS, Fassier J-B, Besen E, Blanck P, Ekberg K, Feuerstein M, et al. Sustaining Work Participation Across the Life Course. Journal of Occupational Rehabilitation. 2016;26(4):465–79.

64. The Society of Occupational Medicine. Long COVID and Return to Work – What Works? [online]. 2022. https://www.som.org.uk/sites/som.org.uk/files/Long_COVID_and_Return_to_Work_What_Works.pdf (accessed 13 May 2024).

65. Ceolta-Smith J, Sparks P, Macniven L, Rayner C, Stanley K. Workers’ experience of long COVID [online]. 2023. https://www.tuc.org.uk/research-analysis/reports/workers-experience-long-covid?page=1#section_header (accessed 13 May 2024).

66. Nastasia I, Rives R. Successful Strategies for Occupational Health and Safety in Small and Medium Enterprises: Insights for a Sustainable Return to Work. Journal of Occupational Rehabilitation. 2024.

67. Skivington K, Matthews L, Simpson SA, Craig P, Baird J, Blazeby JM, et al. A new framework for developing and evaluating complex interventions: update of Medical Research Council guidance. BMJ. 2021;374:n2061.

