## Supplementary Material 1 for "Return to Work with Long COVID: A Rapid Review of Support and Challenges"

### Search Strategy

#### Google Scholar – Search Conducted 27^th^ June 2024

Results = 21

#### Web of Science – Search conducted 27^th^ June 2024

“post covid” or “long covid” or “chronic covid” or “post acute covid” (All fields) AND “return to work” or “sickness absence” or employment or “vocational rehabilitation” or “occupational rehabilitation” or “back to work” or “work participation” or workability or “work ability” or “sick leave” or “illness day*” or “disability leave” or absenteeism (All fields)

Results: **514**

#### OVID – Search conducted 28^th^ June 2024

APA PsycInfo <1806 to June Week 3 2024>

EBM Reviews - Cochrane Central Register of Controlled Trials <May 2024>

EBM Reviews - Cochrane Database of Systematic Reviews <2005 to June 26, 2024>

Embase <1974 to 2024 June 26>

HMIC Health Management Information Consortium <1979 to May 2024>

Ovid MEDLINE(R) and Epub Ahead of Print, In-Process, In-Data-Review & Other Non-Indexed Citations, Daily and Versions <1946 to June 27, 2024>

1	(return to work or sickness absence or employment or vocational rehabilitation or occupational rehabilitation or back to work or work participation or workability or work ability or sick leave or illness day* or disability leave or absenteeism).mp. [mp=ti, ot, ab, hw, kw, tc, id, tm, mf, fx, sh, tx, ct, tn, dm, dv, kf, dq, bt, nm, ox, px, rx, an, ui, sy, ux, mx]	470728

2	(post covid or long covid or chronic covid or post acute covid).mp. [mp=ti, ot, ab, hw, kw, tc, id, tm, mf, fx, sh, tx, ct, tn, dm, dv, kf, dq, bt, nm, ox, px, rx, an, ui, sy, ux, mx]	38426

3	1 and 2	984

4	remove duplicates from 3	**681**
