## Supplementary Material 2 for "Return to Work with Long COVID: A Rapid Review of Support and Challenges"

Supplementary material 2: Study characteristics

| **Study, year of publication** | **Country** | **Study design** | **Population type** | **Sample size *n*** | **Duration of LC symptoms*** | **Percentage female** | **Age of participants** | **Occupation type (if reported)** | **Quality assessment** |
| --- | --- | --- | --- | --- | --- | --- | --- | --- | --- |
| Altmann et al. 2023 (14) | Germany | Prospective cohort | Patients from a cardiac in-patient rehabilitation facility for COVID-19 and LC | 42 (21 COVID-19 & 21 LC) | 2.0 (1-3) months for COVID-19 and 10.1 (4-20) months for LC | 50% (of which, 40% of COVID-19 patients and 60% of LC patients) | LC patients were mean 3 years younger than C19 patients | NR | Medium risk |
| Brehon et al. 2022 (19) | Canada | Retrospective cohort | Workers participating in WCB-Alberta’s Millard Health post-COVID rehabilitation program | 81 | Mean (SD) = 165.2 (73.0) days | 64% | Mean (SD) = 48.9 (10.5) | Health; business, finance, and management; education, law, social, and community government services; trades; other occupations | Low risk |
| Chasco et al. 2022 (25) | USA | Qualitative | Adult patients of the University of Iowa Hospitals and Clinics Post-COVID-19 Clinic | 15 | Average = 11 months | 66.7% | Mean, range = 49.3, 40-68 | NR | Low risk |
| Delgado-Alonso et al. 2022 (12) | Spain | Cross-sectional | Patients with PCS | 77 | Mean (SD) 20.71 (6.50) months | 87.0% | Mean (SD) = 46.31 (7.97) | Occupation by ISCO Classification: Managers, Professionals, Technicians and Associate Professionals, Service and Sales Workers, 8 Plant and Machine Operators, and Assemblers, and Elementary occupations | Low risk |
| Derksen et al. 2023 (34) | Germany | RCT | Patients with PACS. Three cohorts: IG or an ACG and comparison group | 1020 | NR | 74.8% (of which, intervention group: 67.1%, active control group: 73.7%, comparison group: 75.7%) | Mean (SD) of intervention group = 43.47 (12.65), active control group = 44.55 (11.45), comparison group = 46.32 (13.59) | NR | Medium risk |
| Folk 2023 (30) | US, Canada | Qualitative | Academic library employees | 32 | Mean (range) = 16 (1-42) months | 93% | Range = 30-39 - 50–59 | Academic library employees | Medium risk |
| Frisk et al. 2023 (22) | Norway | Quasi-experimental cohort | LC patients | 78 | Mean (SD) = 10.2 (4.8) months | 82% | Range = 18-67 | NR | Low risk |
| Ghali et al. 2023 (21) | France | Cross-sectional | Patients diagnosed with PCS | 86 | Median (range) = 10 (6–13) months | 77.3% | Median (IQR) = 41 (33–48) | NR | Medium risk |
| Gyllensten et al. 2023 (26) | Sweden and Denmark | Qualitative | All were working at least 50 per cent at the time of the recruitment and had long-term effects from Covid that were affecting their work ability | 19 | Median, mean (range) = 18, 15 (4-22) months | 68% | Mean = 54, range = 29-63 | Teacher, web designer, welder,  administrator, psychotherapist, engineer,  manager, cleaner, nurse, caretaker,  assistant nurse, finance assistant | Low risk |
| Kerling et al. 2024 (35) | Germany | RCT | PCS patients | 62 | NR | 68% | Mean (SD) = 46.4 (11.2) | NR | Medium risk |
| Kohn et al. 2022 (27) | Belgium | Qualitative | People with self-reported LC | 97 forum, 33 interviews | *n* (%) = 4–12 weeks: 14 (14); 12 weeks-6 months: 30 (31); > 6 months: 53 (55) | 77% | Range = French speaking: 18->60, Dutch speaking: 25->75 | 20% of population sample worked in health sector | High risk |
| Kvale et al. 2024 (18) | Norway | Prospective cohort | Patients with chronic low back pain, LC, or type 2 diabetes | 241 | NR | 57% | Mean (range) = 48 (19-84) | NR | Medium risk |
| LeGoff et al. 2023 (20) | USA | Retrospective cohort | Primarily frontline and essential workers who were presumed to have been exposed to SARS-CoV-2 during the performance of their duties | 64 | Mean work leave = 11 months | 54.7% (of which, valid group^$^: 42%, invalid group: 64.5%). | Mean = Valid group^$^: 49 years 0 months; Invalid group: 47 years 4 months | Healthcare services; corrections; first responders; facilities maintenance | High risk |
| Lunt et al. 2024 (31) | UK | Qualitative | Female professional workers | 10 | Range = 4 months - >2 years | 100% | Range = 25-63 | NR | Medium risk |
| McNabb et al. 2023 (32) | USA | Qualitative | People with LC | 24 | NR | 54% | Median (IQR) = 46.5 (39.7-55) | 21% of sample population were healthcare workers | Low risk |
| Miller et al. 2024 (33) | UK | Qualitative | Adults with LC | 25 | Mean (SD) = 78 (33) weeks | 68% | Mean (SD, range) = 43.6 (14.7, 19-76) | NR | Low risk |
| Müller et al. 2023 (17) | Germany | Prospective cohort | Healthcare and non-healthcare workers who acquired COVID-19 in the workplace | 127 | Mean, range = 408.81, 124–813 days | 76.40% | Mean (SD) = 50.62 (10.74), range = 21-69 | 70% of population sample worked in health sector | Low risk |
| Nielsen and Yarker 2024 (8) | UK | Qualitative | Workers with LC | 12 | NR | 92% | Average (range) = 45 (30-58) | NR | Low risk |
| Schmachtenberg et al. 2023 (28) | Germany | Qualitative | People with LC by NICE definition | 25 | NR | 72% | Mean = 45, range = 21-67 | Multiple occupation-types including healthcare, management, administration, education and technical work | Low risk |
| Schmid et al. 2024 (16) | Germany | Prospective cohort | Patients with post-COVID fatigue | 64 | Mean (SD) = 11.58 (7.03) months | 70.3% | Mean (SD, range) = 44.1 (11.44, 21-68) | NR | Low risk |
| Stelson et al. 2023 (29) | Multi-national | Qualitative | People living with LC | 510 | Mean, range = 173.8, 134-412 days | 58% | Range = 18 - >60 (80% of participants were 30-60) | NR | Low risk |
| Straßburger et al. 2023 (13) | Germany | Cross-sectional | Post-COVID patients | 184 | Range (% of participants) = 3-6 months (8%); 6-12 months (48%); 12-24 months (27%); >24 months (17%) | 77% | Range = 18->64 | NR | Low risk |
| Tan and Koh 2023 (23) | Singapore | Case study | Participant with LC-type symptoms, but undiagnosed | NA | NA | NA | 27 years | Occupational Health trainee who worked as a government public health officer in an office environment (NA) | Low risk |
| Tanguay et al. 2023 (15) | Canada | Prospective cohort | Participants with LC referred to a telerehabilitation service in the province of Québec | 34 | Mean (SD) = 146.0 (96.0) days | 79% | Mean (SD) = 47.0 (8.8), range = 27-64 | NR | Medium risk |
| Wagner et al. 2022 (24) | Austria | Case study | Female with persistent symptoms post Covid-19 infection | NA | 6.5 months | NA | 55 years | NR | Low risk |

*Can be defined as time from acute COVID-19 infection, duration of LC symptoms or follow-up period, unless stated otherwise. ^$^ Participant group assessed as reporting valid post-COVID-19 symptoms according to the neurocognitive screening evaluation. Definitions: ACG: active control group; IG: intervention group; LC: long COVID; NICE: National Institute for health and Care Excellence; PCS: post-COVID syndrome; PCC: post-COVID condition; SD: standard deviation
