## Supplementary Material 3 for "Return to Work with Long COVID: A Rapid Review of Support and Challenges"

Supplementary material 3: Workplace support for RTW for PwLC

|  |  |  |  |  | **Barriers to support** | | | **Facilitators for support** | |
| --- | --- | --- | --- | --- | --- | --- | --- | --- | --- |
| **Study, year of publication** | **Method** | **Sample size** | **What support?** | **Provider** | **Organisational factors** | | **Individual factors** | **Organisational factors** | **Individual factors** |
| Gyllensten et al. 2023 (26) | Semi-structured focus groups | 19 | Good communication and sufficient support | Manager, peers | Lack of knowledge and understanding of LC |  | |  |  |
|  |  |  | Find a good balance between activity and rest | Self |  |  | |  |  |
|  |  |  | Colleagues share workload | Peers |  |  | |  |  |
|  |  |  | Possibilities to adjust work, e.g. flexible hours, tasks | Management | Adjustments only made short-term |  | |  |  |
| Stelson et al. 2023 (29) | Open ended survey questions | 510 | Remote working | NR |  |  | |  |  |
|  |  |  | Workers' compensation if contracted at work | NR | Lacked documentation of contracting COVID |  | |  |  |
|  |  |  | Medical leave, workplace accommodations and support | NR | Not believed or judged both at work and in medical settings |  | |  |  |
| Schmachtenberg et al. 2023 (28) | Interview | 25 | Emotional support and understanding | Superior, peers | Questioned or doubted by supervisors | Role conflicts and identity crises | |  |  |
|  |  |  | Home office (WFH) | Workplace | Not recognising limitations | Limited contact with colleagues or customers | |  |  |
|  |  |  | Vocational reintegration program |  |  |  | |  |  |
|  |  |  | Reduce working hours |  |  | Decrease income | |  |  |
| Tan and Koh 2023 (23) | Case study | 1 | Self-developed rehabilitation program: gradual fitness plan for 2 months | Self | No occupational health service provision |  | |  |  |
| Chasco et al. 2022 (25) | Semi-structured phone interviews | 15 | Checking in with the participant, approving SL or vacation time, accommodating medical appointments, adapting work tasks | Employer | Colleagues and employers’ perception of participants’ ability to work; Stigma at Work | Self-perception of ability to work, related to LC symptoms | | Recognition of PACS as a medical condition; Eligibility for disability benefits; Policy action | Take a pragmatic approach to symptom management: not hiding symptoms |
| Delgado-Alonso et al. 2022 (12) | Survey | 38 | Reduced working hours. Job adaptation, e.g. more breaks, telework, cognitive aids, or a position change. | NR | Some reported not possible to make needed adaptations |  | |  |  |
| Kohn et al. 2022 (27) | Semi-structured online interviews, forum | 97 forum, 31 interviews | ‘Medical part-time work’ | PS | Administrative burden | Income loss | |  |  |
|  |  |  | Adjust the rhythm: taking breaks |  |  | Psychological strain, feelings of guilt | |  |  |
|  |  |  | Teleworking |  |  |  | |  |  |
|  |  |  | Adapting work, in consultation with occupational physician |  |  |  | | Recognition of LC and C19 as ODs |  |
| Folk 2023 (30) | Online survey free-text questions | 32 | Supervisors were willing to be flexible | Supervisors | A lack of flexibility |  | |  |  |
|  |  |  | Colleagues expressed concern and validation of LC. Willing to help | Colleagues | A demonstrated lack of understanding about what they were experiencing |  | |  |  |
| Lunt et al. 2024 (31) | Online semi-structured interviews | 10 | Managerial/peer support: Management and HR consultations | Workplace | Excluded LC employees | A sense of loss over career aspirations | | Empathic line management | Communication strategies |
|  |  |  |  |  | Colleagues' insensitive attitudes |  | | Peer support that accepts conditions |  |
|  |  |  | Organisational support/ OH: Individualised RTW planning | Workplace | Internal OH policing |  | | Organisation policies that provide certainty of support and income | Knowing your employment rights |
|  |  |  |  |  | Standardised OH absence management insensitive to individual needs |  | | Encouraging organisational cultures; Education on how to support LC |  |
| McNabb et al. 2023 (32) | Interview | 24 | Remote working | Employer | Chronic illness needs were misunderstood by employers | Stress of not meeting previous productivity | |  |  |
| Miller et al. 2024 (33) | Interview | 25 | Opportunities for rest and breaks; Phased RTW program, hybrid working | PS | Misunderstanding from colleagues leading to friction and hostility | Stress over conflict at work; Reduced income | |  |  |
|  |  |  | Workplace adjustments: quiet area | PS | Unable to provide medical verification | Feelings of failure and loss of identity | |  |  |
|  |  |  | Mandatory training for employers on chronic conditions | PS |  |  | |  |  |
|  |  |  | Access to occupational therapist | PS |  |  | |  |  |
|  |  |  | LC to be recognised as a disability to industrial injury if acquired at work | PS |  |  | |  |  |
| Nielsen & Yarker 2024 (8) | Interview | 12 | Emotional and instrumental support, e.g. work adjustments | Colleagues | Lack of understanding or toxic work environment e.g. alienation | Fluctuating symptoms hindered work confidence | | Colleagues' acceptance of their reduced work functioning |  |
|  |  |  | LC staff support groups to share experiences and practical advice |  |  |  | | Support groups empower advocacy within organisations |  |
|  |  |  | Necessary work adjustments | Manager | Line managers lack understanding of LC | Concerns about limited cognitive functioning | | Line managers understanding |  |
|  |  |  |  |  | Pressure from managers to ignore symptoms | Questioned whether working was beneficial | |  |  |
|  |  |  | Part-time work | Manager | Pressure to go part-time | Leading to income loss | |  |  |
|  |  |  | OH and HR support: phased RTW | Organisation | Lack of understanding in OH about mental or neurocognitive aspects of LC | Occupational identity struggle, feelings of guilt | | OH recognised their needs and baseline capabilities, tailored support | Redefined occupational identity |
|  |  |  |  |  | HR phased RTW policies inflexible, not suited to LC |  | |  | Participants accepting a LC sufferer identity |
|  |  |  |  |  | Disconnect between OH and HR |  | |  |  |
|  |  |  |  |  | HR failed to address inappropriate manager actions |  | |  |  |
|  |  |  | Wider support | Government | Lack of government policies addressing fluctuation of LC | Lack of recognition from broader society | |  |  |
|  |  |  |  |  | Lack of knowledge about how to support PwLC |  | |  |  |
| Straßburger et al. 2023 (13) | Survey | 184 | Reduce working hours; occupational reintegration plan; adjustments to the workplace; adjustment of tasks; health courses; general consultation; job coaching | Workplace |  |  | | Structural changes e.g. modification of workplace, working hours and tasks |  |

Abbreviations: C19: Covid-19, SARS-CoV-2 infection, SARS-CoV-2 positive; LC: Long COVID; RTW: return, returning or returned to work; PACS: post-acute COVID-19 syndrome; OD: Occupational disease or illness; OH: Occupational health; PS: participants suggestions; pwLC: people with long covid
