## Supplementary Material 4 for "Return to Work with Long COVID: A Rapid Review of Support and Challenges"

Supplementary material 4: Interventions supporting RTW with LC

| Study | What intervention? | Who provided it? | Participants | Sample size | Outcome measures | Unit of measurement | Results | P value |
| --- | --- | --- | --- | --- | --- | --- | --- | --- |
| Derksen et al 2023 (34) | Interdisciplinary health care facilitation program based on professional personal pilot support and digital interventions, both delivered as medical internet aid. | University researchers, funded by German Bavarian State | Patients with PACS in three groups: Intervention (IG), active control (ACG), comparison (CompG) | 1020 (IG=73, ACG=152, CompG= 795) | WAI | Mean (SD) | IG: T2 5.13 (2.56), T4 5.43 (2.40), T5 5.69 (2.87); ACG: T2 4.97 (2.21), T4 5.65 (2.51), T5 5.65 (2.77); CompG: T2 4.46 (2.65), T4 4.72 (2.89), T5 N/A |  |
|  |  |  |  |  | Effects Between IG and ACG vs CompG | Estimated coefficient (SE) | T2 vs T4: 0.28 (0.15), p=.063; IG and ACG vs CompG: 0.78 (0.29), p=.007; |  |
|  |  |  |  |  | Effects Between IG and ACG |  | T2 vs T4: -0.58 (0.13), p<.001; T4 vs T5:-0.34 (0.13), p=.010; IG vs ACG: -0.07 (0.43), p=.875 |  |
| LeGoff et al 2023 (20) | A Neurocognitive screening evaluation (NCSE) for assessment and treatment to facilitate employee recovery and RTW | Outpatient mental health provider panel (US Workers Compensation) | California employees with PCC claims due to 'brain fog' | 64 | LWDs | Mean(SD), Median, Range | Before referral: 244.1 (126.4), 221.4 , 49-536; Post NCSE (from the date of the NCSE to the treating psychologist’s RTW release date on either modified or full duty): 67.4 (127.7), 19.5, 1-286. P < 0.001 | P < 0.001 |
| Frisk et al 2023 (22) | Clinical intervention consisted of three phases: Pre‑treatment preparation, concentrated micro‑choice based rehabilitation and Integrating changes into everyday living | Haukeland University Hospital (Bergen, Norway) | LC patients | 78 | SL | % of employed participants on SL | 63% at baseline to 43% at 3-month (OR = 0.2, p = 0.02) | p = 0.02 |
| Tanguay et al 2023 (15) | Telerehabilitation based on the STOP-REST-PACE approach and occupational therapy | Rehabilitation service at Université de Sherbrooke (Canada) | Participants with LC | 30 | Medical leave, working PT or FT | n | Week 0: 20, 6, 4; Week 8: 17, 11, 2; Week 12: 17, 9, 4 | NA |
| Altmann et al 2023 (14) | Rehabilitation program included multimodal respiratory therapy, endurance and resistance muscular training, psychological assistance, and education | Cardiac in-patient rehabilitation, by German Pension Fund and German Workers Accidents Insurance | C19 cases acquired during professional duty | 21 (LC) | Fit/Unfit for work | % of Fit for work at discharge, RTW in 6 months, Unfit for work after 6 months | 33, 52, 29 (patients admitted mean 10.1 months after C19 infection) |  |
|  |  |  |  | 21 (C19) |  |  | 5, 95, 5 (patients admitted mean 2.0 months after C19 infection) |  |
| Müller et al 2023 (17) | Inpatient multidisciplinary rehabilitation program included medical treatment and care, physical and psychological treatments | Rehabilitation in a Hospital, registered by accident insurance providers (Germany) | Workplace acquired C19 with PAC as a recognized OD or accident | 115 | WAI | Median Score (IQR) | Pre rehabilitation: 24.75 (21–28); Post rehabilitation: 24.75 (21–28), p = 0.408 | p = 0.408 |
| Ghali et al 2023 (21) | PCS patients were given a leaflet about pacing strategies and asked to keep a diary | University Hospital (France) | Patients diagnosed with PCS | 86 | RTW (Recovery) | %, OR [95% CI] | 29 (33.7%) experienced recovery and RTW, of which 10 (34.5%) returned to FT work, 19 (65.5%) returned to PT work. 20 (23.3%) showed improvement in their health status but not RTW, and 37 (43%) did not recover or improved. Association between adherence to pacing strategies (EPS) and RTW is siginificant, OR= 40.43 [95% CI 6.22–262.64], p < 0.001 | p < 0.001 |
| Brehon et al 2022 (19) | Multidisciplinary program consists of occupational, physical, and exercise therapy along with psychology, nursing, and medical interventions, including psychoeducational, guidance on pacing and breathing strategies | Workers’ Compensation Board of Alberta (Canada) | LC claims due to workplace exposure | 81 | RTW | n (%) | 12% Working at admission, 53% RTW at discharge |  |
|  |  |  | Of those RTW at discharge | 43 | RTW | n (%) | 7% Returned to regular duties, 93% Returned to modified duties |  |
| Wagner et al 2022 (24) | Pulsed electromagnetic field (PEMF) treatment for post-covid fatigue | Outpatient clinic (Austria) | A 55-year-old female | 1 | WAI short version | Score | WAI before treatment 21.5 (critical); WAI immediately after treatment 40 (good); WAI 6 weeks after treatment 40 (good) | NA |
| Kerling et al 2024 (35) | 3-month home exercise plan combining moderate and intense physical activities per week | Funded research: Hannover Medical School | Post-COVID-19 syndrome (PCS) patients | 62 | WAI | Score 7- 49 for 7 questions | Work ability did not change in either group with the intervention. IG (n=30): change from baseline – 0.3 ± 5.7 (cohens d 0.05); CG (n=32): 0.7 ± 3.5 (cohens d – 0.19); Mean difference between groups over time [CI95%] 1.0 [– 1.9; 3.8] | NR |
| Kvale et al 2024 (18) | Three phases: (1) preparing for change, (2) the concentrated intervention for 3–4 days, and (3) integrating change into everyday life. Patients were taught and practiced how to monitor and target seemingly insignificant everyday micro-choices to break behavioural patterns contributed to health problems. | Funded research: Haukeland University Hospital | Patients with chronic low back pain, long COVID, or type 2 diabetes | 104 low back pain, 76 LC, and 61 type-2 diabetes | Work and Social Adjustment Scale (WSAS) | Score 0 (not at all) to 8 (very severely), higher scores indicating higher impairment | The mixed regressions analyses showed that level of functioning measured by WSAS improved at follow-up for chronic low back pain (b = −4.06, Z = 5.19, P < .001) and long COVID patients (b = −7.66, Z = 8.37, P < .001), but not for diabetes patients (b = −1.88, Z = 1.58, p = .11). | LC P < .001 |
| Schmid et al 2024 | 14-day multimodal integrative inpatient program, consists of individual therapeutic modalities: vitamins/ nutrients, hydrotherapy, thermotherapy, mind–body medicine, phytotherapy, exercise therapy, naturopathy, nutrition, and other therapies | Clinic for Internal and Integrative Medicine in Bamberg, Germany | Patients with post-COVID fatigue | 39 | Self-reported ability to work | 11-point scale | T1 (admission) vs T2 (discharge) vs T3 (6 months after discharge): Self-reported work ability increased significantly from immediately before the 14-day treatment (T1: M=2.54, SD=2.23; T2: M=4.26, SD=2.60; and T3: M=4.41, SD=3.23) | p<0.001 for T1 vs T2 and T1 vs T3 |

Abbreviations: C19: Covid-19, SARS-CoV-2 infection, SARS-CoV-2 positive; LC: Long Covid; WAI: work ability index; RTW: return, returning or returned to work; PACS: postacute COVID-19 syndrome; LWDs: Lost workdays; SL: Sick leave; PT: Part-time; FT: Full-time; PAC: post-acute COVID-19; OD: occupational disease
